# Subclinical ocular inflammation in persons recovered from ambulatory COVID-19

**DOI:** 10.1101/2020.09.22.20128140

**Authors:** Mathieu F. Bakhoum, Michele Ritter, Anupam K. Garg, Alison X Chan, Christine Y. Bakhoum, Davey M. Smith

## Abstract

Coronavirus disease 2019 (COVID-19) is characterized by striking variability in clinical severity, and a hyperinflammatory response in the lung is associated with high mortality. Little is known about the extent and duration of inflammation in persons recovering from COVID-19. Here, we used spectral domain optical coherence tomography (SD-OCT) to detect the presence of inflammatory cells in the vitreous cavity, an immune-privileged microenvironment, in persons recovered from COVID-19. Our results provide quasi-histologic evidence that neuroinflammation is present in persons who recovered from COVID-19, only one of whom required hospitalization. Our results also suggest that persons who feel that their recovery is incomplete have evidence of subclinical eye inflammation, which may be a marker of residual inflammation elsewhere as well.

## INTRODUCTION

Coronavirus disease 2019 (COVID-19) is characterized by striking variability in clinical severity, and a hyperinflammatory response in the lung is associated with high mortality.[1, 2] Little is known about the extent and duration of inflammation in persons recovering from COVID-19. Here, we used spectral domain optical coherence tomography (SD-OCT) to image individual cells in the vitreous cavity, an immune-privileged microenvironment, in persons recovered from COVID-19.

## METHODS

Individuals with history of COVID-19 but no history of uveitis were recruited. A 97-raster SD-OCT scan (Spectralis, Heidelberg) was obtained. Transverse B-scans were reviewed for the presence of cells in the vitreous cavity.[3] Grading of cells in the subhyaloid space was defined as ‘*rare*’ < 50 cells, *‘few’* 50 to 194 cells and *‘many* >194 cells (average of more than 1 cell per B-scan). In eyes where a subhyaloid space was not present, ‘many cells’ were defined as >776 hyperreflective foci (average of more than 4 cells per B-scan).

## RESULTS

Fifteen participants were evaluated (6 women and 9 men). All but one were previously diagnosed with active SARS-CoV-2 infection by polymerase chain reaction (PCR) 58 to 85 days prior to evaluation. One participant had symptoms consistent with COVID-19 and a simultaneous household contact with confirmed COVID-19. Respiratory symptoms were present in all participants (Table). Only Participant 14 required hospitalization. All met the criteria of recovery from the active viral phase.[4] Time from recovery ranged from 34 to 60 days (median 50 days). Post-recovery, four persons reported cough, two reported lower extremity weakness and numbness, one reported headaches, and one reported photophobia. On SD-OCT scans, 4 persons had few cells and 3 had many cells (**Table** and **Figure**). Participants 4 and 14 had SD-OCT scans at our institution 6 months and 2 years prior that showed none to rare cells (**Figure**, *panels **C** and **D***), indicating that inflammation occurred recently. Participants were asked at the time of examination whether they felt they had a complete recovery. Eleven persons perceived that they are back to their baseline (median 50 days since recovery), while 4 did not (median 47 days since recovery). Of those who felt they had a complete recovery, 8 had rare cells; while among those who had residual symptoms 2 had few cells and 2 had many cells. The grading scheme was applied to OCT scans from 14 individuals with no history of COVID; 13 had rare cells and 1 had few cells (p = 0.006) (**Table** and **Figure**).

**Figure.**
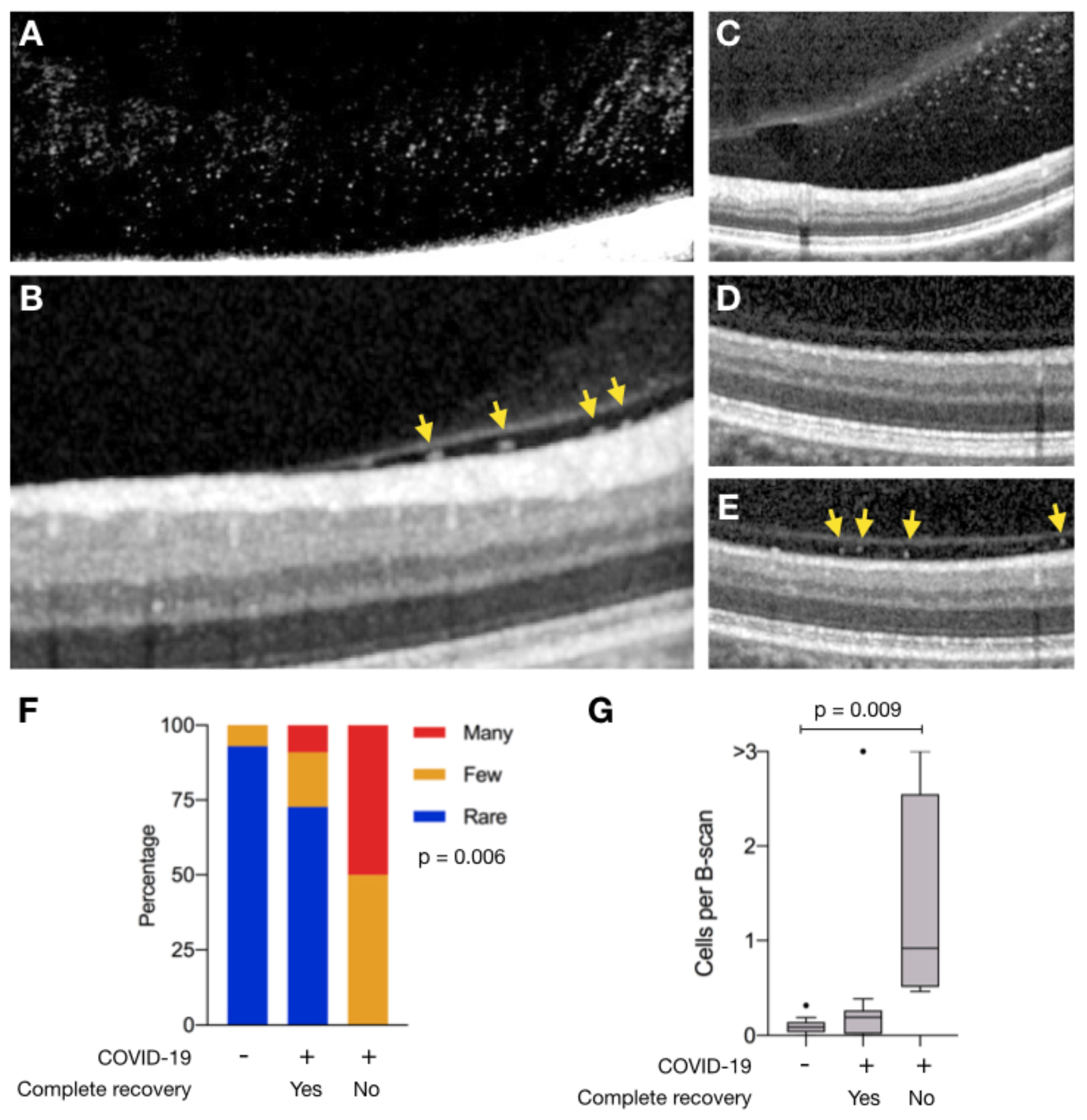
Vitreous cells in persons recovered from COVID-19. SD-OCT transverse sections of A) the posterior vitreous near the inferior macular vascular vessels in Participant 1 with many vitreous cells, B) the retina vitreous interface demonstrating cells (yellow arrows) in the sub-hyaloid space in Participant 5 and C) Participant 4. Identical SD-OCT transverse sections of the retina and vitreous of the right eye demonstrating many cells in the subhyaloid space after recovering from COVID-19 (D), and no vitreous cells 2 years prior (E). Corresponding infrared *en face* images of the retina (not shown) were used to determine the exact reference location. Image in panel A was digitally enhanced to highlight vitreous cells. The adjustment has been made to the entire image. (F) The distribution of vitreous cells grading (rare, blue; few, orange; many, red) among individuals with no history of COVID-19, and those with prior COVID-19 infection who fully recovered (Yes) and those who did not (No). Significance was determined using Pearson’s chi-squared test (p = 0.006). (G) Box plots showing the number of vitreous cells per transverse B-scans among individuals with no history of COVID-19, and those with prior COVID-19 infection who fully recovered (Yes) and those who did not (No). Significance was determined using one-way ANOVA (p = 0.01) and student’s t test with Tukey’s correction.

**Table.** Demographics and clinical characteristics of study participants. Days since SARS-CoV2 were estimated based on onset of symptoms or a positive nucleic acid test, whichever is earlier. The duration of illness was defined as time from onset of symptoms to cure based on either negative SARS-CoV-2 PCR test or complete resolution of symptoms for at least 3 days. Negative PCR tests were obtained via nasal or oropharyngeal swabs; NT, no repeat testing. Systemic symptoms reported after recovery from COVID-19. The use of inhaler or systemic steroid medications is noted. Participants were asked if they had a complete symptomatic recovery. Vitreous cells grading as defined in Methods. Grading of vitreous cells in Participant 1 was based on hyperreflective foci in the vitreous cortex, as there was no subhyaloid space.

|  | Age group, gender | Symptoms | Days since COVID-19 onset | Days since recovery | Method of negative PCR | Symptoms post-recovery | Steroid medications | Participant reporting symptomatic recovery | Vitreous cells grading |
| --- | --- | --- | --- | --- | --- | --- | --- | --- | --- |
| 1 | 30-40, M | Fever, cough, chest pain, fatigue, anosmia, ageusia | 69 | 35 | Nasal, oropharyngeal | Headaches, Cough, photophobia | Steroid inhaler | No | Many |
| 2 | 30-40, M | Fever, cough, anosmia, ageusia | 70 | 40 | NT | Lower extremity weakness/numbness | None | Yes | Rare |
| 3 | 40-50, M | Pharyngitis, cough, anosmia | 60 | 34 | Nasal x2 | None | None | Yes | Rare |
| 4 | 40-50, F | Fever, cough, pharyngitis, rhinorrhea | 68 | 50 | NT | Cough | Steroid inhaler | Yes | Many |
| 5 | 40-50,<br>F | Sinus congestion,<br>dyspnea | 71 | 50 | Oropharyngeal | None | Steroid inhaler | No | Few |
| 6 | 50-60,<br>M | Cough, Fever | 85 | 59 | Nasal x2 | Lower extremity<br>weakness/numbness | None | No | Many |
| 7 | 50-60,<br>M | Cough, Fatigue | 73 | 53 | NT | None | None | Yes | Rare |
| 8 | 50-60,<br>F | Fever, otalgia<br>headache, fatigue | 58 | 45 | Nasal x2 | None | Steroid inhaler | Yes | Few |
| 9 | 60-70,<br>M | Fever, cough,<br>fatigue, anosmia,<br>ageusia | 65 | 45 | NT | None | None | Yes | Rare |
| 10 | 70-80,<br>F | Chest pain, sinus<br>congestion,<br>headache | 73 | 58 | Nasal x2 | None | None | Yes | Rare |
| 11 | 70-80,<br>M | Cough, fever, body<br>aches | 59 | 51 | Nasal | Post-nasal drip, dry<br>cough | Systemic<br>steroid | Yes | Rare |
| 12 | 50-60,<br>F | Fever, cough,<br>Fatigue, anosmia,<br>ageusia | 71 | 47 | Nasal x2 | None | None | Yes | Few |
| 13 | 20-30,<br>F | Fever, cough,<br>rhinorrhea | 60 | 56 | NT | None | None | Yes | Rare |
| 14 | 60-70,<br>M | Fever, cough and<br>respiratory failure | 85 | 43 | Nasal x2 | Cough | Inhaled | No | Few |
| 15 | 60-70,<br>M | Fever, cough | 74 | 60 | NT | None | None | Yes | Rare |

## DISCUSSION

Here, we provide **quasi-histologic evidence that neuroinflammation is present in persons who recovered from COVID-19, only one of whom required hospitalization**. On OCT imaging, presence of numerous cells in the vitreous cavity is abnormal. Cells in the vitreous result from inflammation, hemorrhage or a neoplastic process.[3, 5, 6] Strikingly, in two participants OCT scans prior to their COVID-19 illness showed no evidence of inflammation in the vitreous, while their scans after COVID-19 showed hyperreflective foci, strongly suggesting that even ambulatory COVID-19 may lead to inflammatory cells persisting in the eye up to 1 month after recovery. Persons who felt that their recovery was incomplete had more cells, which likely suggests residual inflammation elsewhere. Our findings are congruent with emerging reports demonstrating that recovery from COVID-19 may be complicated by post-viral inflammation, and full symptomatic recovery may not occur until weeks after a positive test result, even in younger individuals [7, 8].

**This study has limitations**. The identification of inflammatory cells relied on SD-OCT imaging, and there were no molecular or cellular analysis of vitreous biopsies. The specificity of this finding to COVID-19 is unknown, as no similar study to our knowledge has been conducted in persons recovering from other respiratory viral or systemic illnesses. A larger study is needed to determine the true prevalence of vitreous cells in persons recovering from COVID-19, and a longitudinal study is needed to determine its long-term ocular health sequelae.

## Data Availability

N/A

## Conflict of interest

The authors declare no conflict of interest.

## Notes

### Competing Interest Statement

The authors have declared no competing interest.

### Author Declarations

IRB was obtained from the University of California San Diego

### Summary of Updates

1. Typo in legend - 'COVID-10', changed to COVID-19 2. Panels D and E were swapped

## References

1. Zhang, X., et al., Viral and host factors related to the clinical outcome of COVID-19. Nature, 2020.

2. Huang, C., et al., Clinical features of patients infected with 2019 novel coronavirus in Wuhan, China. Lancet, 2020. 395(10223): p. 497-506.

3. Saito, M., I.A. Barbazetto, and R.F. Spaide, Intravitreal cellular infiltrate imaged as punctate spots by spectral-domain optical coherence tomography in eyes with posterior segment inflammatory disease. Retina, 2013. 33(3): p. 559-65.

4. Caberlotto, L. Symptom-Based Strategy to Discontinue Isolation for Persons with COVID-19. 2020 May 23, 2012]; Available from: https://www.cdc.gov/coronavirus/2019-ncov/community/strategy-discontinue-isolation.html.

5. Pichi, F., et al., Optical coherence tomography diagnostic signs in posterior uveitis. Prog Retin Eye Res, 2020. 75: p. 100797.

6. Zhao, H., et al., Longitudinal observation of OCT imaging is a valuable tool to monitor primary vitreoretinal lymphoma treated with intravitreal injections of methotrexate. BMC Ophthalmol, 2020. 20(1): p. 10.

7. Puntmann, V.O., et al., Outcomes of Cardiovascular Magnetic Resonance Imaging in Patients Recently Recovered From Coronavirus Disease 2019 (COVID-19). JAMA Cardiol, 2020.

8. Tenforde, M.W., et al., Symptom Duration and Risk Factors for Delayed Return to Usual Health Among Outpatients with COVID-19 in a Multistate Health Care Systems Network - United States, March-June 2020. MMWR Morb Mortal Wkly Rep, 2020. 69(30): p. 993-998.

